# From multiplex serology to serolomics – a novel approach to the antibody response against the SARS-CoV-2 proteome

**DOI:** 10.1101/2020.10.19.20214916

**Authors:** Julia Butt, Rajagopal Murugan, Theresa Hippchen, Sylvia Olberg, Monique van Straaten, Hedda Wardemann, Erec Stebbins, Hans-Georg Kräusslich, Ralf Bartenschlager, Hermann Brenner, Vibor Laketa, Ben Schöttker, Barbara Müller, Uta Merle, Tim Waterboer

## Abstract

**Background:** The emerging SARS-CoV-2 pandemic entails an urgent need for specific and sensitive high-throughput serological assays to assess SARS-CoV-2 epidemiology. We therefore aimed at developing a fluorescent-bead based SARS-CoV-2 multiplex serology assay for detection of antibody responses to the SARS-CoV-2 proteome.

**Methods:** Proteins of the SARS-CoV-2 proteome and protein N of SARS-CoV-1 and common cold Coronaviruses (ccCoVs) were recombinantly expressed in *E. coli* or HEK293 cells. Assay performance was assessed in a Covid-19 case cohort (n=48 hospitalized patients from Heidelberg) as well as n=85 age- and sex-matched pre-pandemic controls from the ESTHER study. Assay validation included comparison with home-made immunofluorescence and commercial Enzyme-linked immunosorbent (ELISA) assays.

**Results:** A sensitivity of 100% (95% CI: 86%-100%) was achieved in Covid-19 patients 14 days post symptom onset with dual sero-positivity to SARS-CoV-2 N and the receptor-binding domain of the spike protein. The specificity obtained with this algorithm was 100% (95% CI: 96%-100%). Antibody responses to ccCoVs N were abundantly high and did not correlate with those to SARS-CoV-2 N. Inclusion of additional SARS-CoV-2 proteins as well as separate assessment of immunoglobulin (Ig) classes M, A, and G allowed for explorative analyses regarding disease progression and course of antibody response.

**Conclusion:** This newly developed SARS-CoV-2 multiplex serology assay achieved high sensitivity and specificity to determine SARS-CoV-2 sero-positivity. Its high throughput ability allows epidemiologic SARS-CoV-2 research in large population-based studies. Inclusion of additional pathogens into the panel as well as separate assessment of Ig isotypes will furthermore allow addressing research questions beyond SARS-CoV-2 sero-prevalence.

## INTRODUCTION

The SARS-CoV-2 pandemic has emerged worldwide, but there is still a lack of knowledge on the epidemiology of infection. Large-scale population-based studies would not only provide reliable prevalence estimates but also identify factors associated with the infection and transmission. Consequently, there is an urgent need for assays that provide high-throughput methodology. Direct detection of the infectious SARS-CoV-2 or its RNA genome is limited to a specific time frame after infection and only provides information about current but not past infections. In contrast, antibody responses indicate current and past infections and allow for a cross-sectional assessment of SARS-CoV-2 cumulative exposure in a given population. Current serological assays are mostly ELISA- or (electro)chemiluminescence-based and limited to a single antigen, either the nucleocapsid protein (N) or the spike protein (S) (Post et al., 2020). Often, subdomains of S are used as antigens, including the N-terminal S1 domain, which is cleaved from the C-terminal S2 domain during host cell attachment and entry, and the receptor-binding domain (RBD) as part of the S1 domain (Post et al., 2020). Both, S1 and S1-RBD are highly glycosylated, which is important for correct conformation of the protein (Walls et al., 2020). Proteins N and S share high sequence homologies to their counterparts of other Coronaviruses (CoVs), including SARS-CoV-1, and endemic common cold CoVs (ccCoVs) NL63, 229E, HKU1, and OC43, potentially resulting in cross-reactive antibody responses and consequently lowered specificity (Meyer et al., 2014). A multiplex approach analyzing multiple antigens in a time- and labor-efficient manner would potentially increase specificity of detecting current and/or past SARS-CoV-2 exposure, and allow identifying antibody patterns meaningful for e.g. prediction of disease course.

Thus far, only a few studies have employed either microarray or fluorescent-bead based technologies to develop multiplex SARS-CoV-2 serological assays (Becker et al., 2020; Dobaño et al., 2020; Jiang et al., 2020; Roxhed et al., 2020; Zamecnik et al., 2020; Zhang et al., 2020), all providing high specificity and sensitivity in detecting SARS-CoV-2 antibodies by varying combinations of proteins N and S, as well as subdomains or peptides thereof. Microarray-based studies utilized peptides or proteins of the SARS-CoV-2 proteome (Jiang et al., 2020; Zamecnik et al., 2020; Zhang et al., 2020) allowing for assessment of the immunogenicity of proteins other than N and S. In contrast to fluorescent-bead based technologies, microarray-based assays are, however, not suited for high-throughput analyses of large sample sets.

Here, we report the development of a fluorescent-bead based SARS-CoV-2 multiplex serology assay for the detection of antibody responses to the SARS-CoV-2 proteome, including proteins N and S, either in full-length or as their respective subdomains N-EP3 (a predicted B-cell epitope of protein N (Grifoni et al., 2020)) and S1, S1-RBD, S2, and a shorter fragment S2’ (Hoffmann et al., 2020; Stadlbauer et al., 2020). This set-up will potentially allow achieving an exceptionally high specificity and sensitivity by combined antigen algorithms for SARS-CoV-2 sero-positivity. In addition, we aimed to include proteins of the entire SARS-CoV-2 proteome to allow for association studies beyond mere sero-prevalence, as well as the N proteins of related CoVs to assess potential cross-reactive antibody responses. We furthermore aimed for performing assay validation against multiple gold-standard assays in a well-characterized local case cohort of Covid-19 patients as well as age- and sex-matched pre-pandemic controls.

## METHODS

### Study Population

The cohort of Covid-19 cases included in this study was recruited at the Heidelberg University Clinics (Heidelberg, Germany) between March, 18^th^, and May, 22^nd^, 2020. Presence of SARS-CoV-2 infection was confirmed by RT-PCR as described previously (Dao Thi et al., 2020). Briefly, RNA was isolated from nasopharyngeal swabs using QIAGEN Kits (QIAGEN, Hilden, Germany) automated on the QIASymphony instrument (DSP Virus pathogen mini kits). Extracted samples were used in a RT-PCR reaction, carried out using various reagent mixes: LightMix Modular SARS and Wuhan CoV E-gene, LightMix Modular SARS and Wuhan CoV N-gene, LightMix Modular Wuhan CoV RdRP-gene and LightMix Modular EAV RNA Extraction Control (TIBMOLBIOL, Berlin, Germany) and LightCycler Multiplex RNA Virus Master (Roche, Mannheim, Germany) according to manufacturer’s instructions. RT-PCR was performed on LightCycler 480 instruments (Roche, Mannheim, Germany). Tests were performed and interpreted according to the manufacturer’s instructions. Participants provided information on age, sex, and Covid-19 symptoms, if any, as well as date of symptom onset. The need and type of oxygenation as well as death due to Covid-19 in hospitalized patients was documented. Blood samples were drawn from each participant at baseline and in the follow-up in varying time intervals dependent on whether the patient was hospitalized and the duration of hospital stay. In total, n=192 serum samples were shipped to the German Cancer Research Center (DKFZ, Heidelberg, Germany) for serological analyses. Of these, n=18 samples had to be excluded due to mis-labelling of the sample tube, technical errors that had occurred during the multiplex serology analysis, a negative SARS-CoV-2 PCR test, or missing information on onset of symptoms. Thus, the final serum set included in total n=174 sera. N=156 serum samples originated from n=48 hospitalized Covid-19 patients, and n=18 serum samples from n=15 Covid-19 patients who experienced only mild symptoms and were not hospitalized. The median age among hospitalized patients was 62 years (range 23 to 85 years) and 33% of hospitalized patients were female. Among non-hospitalized patients, the median age was 54 years (range 27 to 70 years) and 60% were female.

The study was approved by the Ethics Committee of the Medical Faculty Heidelberg (approval number S-148/2020) and conducted in accordance with the Declaration of Helsinki.

Serum samples from pre-pandemic controls were obtained from the ESTHER I study (“Epidemiologische Studie zu Chancen der Verhütung, Früherkennung und optimierten Therapie chronischer Erkrankungen in der älteren Bevölkerung”), a prospective cohort that enrolled in total 9,940 subjects aged 50 to 75 years between July 2000 and December 2002 in Saarland, Germany. Participants were recruited by their general practitioner during a general health check‐up examination and provided a blood sample as well as sociodemographic information (Brenner et al., 2007). We randomly selected n=88 participants to frequency match the age- and sex-distribution among the above-described hospitalized n=48 Covid-19 patients. After multiplex serology analysis, n=3 serum samples were excluded due to technical errors. The median age among the remaining n=85 pre-pandemic controls was 62 years (range 50 to 75 years) and 33% were female.

The ESTHER study was approved by the ethics committees of the University of Heidelberg (approval number S-058/2000) and the medical board of the state of Saarland (approval number 67/00). Written informed consent was obtained from each participant.

### Selection and Recombinant expression of SARS-CoV-2 Proteins

For the development of SARS-CoV-2 multiplex serology we attempted to express the near-complete SARS-CoV-2 proteome (NCBI accession no. NC_045512.2) (**Table 1**) (Wu et al., 2020). Non-structural proteins (NSP) 3, 4, and 6 were not included in the antigen panel, since these proteins were predicted to have multiple hydrophobic regions posing challenges to efficient expression. We furthermore excluded NSP11 because this protein is composed of only 11 amino acids. Since the SARS-CoV-2 spike (S) protein is cleaved by host furin, we included the respective fragments S1 and S2, as well as an additional shorter fragment of S2, i.e. S2’ (Hoffmann et al., 2020). Proteins N and S1 were previously used in serological analyses (Post et al., 2020). We additionally included SARS-CoV-2 sub-fragments (N-EP3 and S1-RBD) that might induce less cross-reactive antibody responses to related Coronaviruses (Grifoni et al., 2020). All proteins were recombinantly expressed as fusion proteins in *E. coli* BL21 with an N-terminal glutathione S-transferase (GST) and a C-terminal tag consisting of the 11 C-terminal amino acid residues of the large T antigen of simian virus 40, as described previously (Waterboer et al., 2005). Gene synthesis and subcloning in the respective vector pGEX4T3tag was performed by Eurofins (Ebersberg, Germany). All recombinantly expressed proteins underwent quality controls to confirm the correctness of the construct (DNA extraction and PCR with subsequent sequence analysis) as well as sufficient expression yields of full-length protein (semi-quantitative ELISA to detect the C-terminal tag sequence and Western Blots against both terminal tags). All proteins but NSP12 and NSP13 passed the quality controls. These two proteins were therefore excluded from further analyses.

**Table 1:** Selected proteins of SARS-CoV-2 and additional Coronaviruses for multiplex serology.

| Antigen | Accession No. | Selected AA | Expression system | N-terminal tag | Cut-off [MFI]<sup>b</sup> |
| --- | --- | --- | --- | --- | --- |
| **SARS-CoV-2 (NC_045512.2)** |  |  |  |  |  |
| NSP1 | YP_009725297.1 | 1-180 | *E. coli* BL21 | GST | 1107 |
| NSP2 | YP_009725298.1 | 1-638 | *E. coli* BL21 | GST | 1391 |
| NSP5 | YP_009725301.1 | 1-306 | *E. coli* BL21 | GST | 1044 |
| NSP7 | YP_009725303.1 | 1-83 | *E. coli* BL21 | GST | 1719 |
| NSP8 | YP_009725304.1 | 1-198 | *E. coli* BL21 | GST | 849 |
| NSP9 | YP_009725305.1 | 1-113 | *E. coli* BL21 | GST | 605 |
| NSP10 | YP_009725306.1 | 1-139 | *E. coli* BL21 | GST | 147 |
| NSP12 | YP_009725307.1 | 1-932 | *E. coli* BL21 | GST | nd |
| NSP13 | YP_009725308.1 | 1-601 | *E. coli* BL21 | GST | nd |
| NSP14 | YP_009725309.1 | 1-527 | *E. coli* BL21 | GST | 106 |
| NSP15 | YP_009725310.1 | 1-346 | *E. coli* BL21 | GST | 135 |
| NSP16 | YP_009725311.1 | 1-298 | *E. coli* BL21 | GST | 387 |
| S1 | YP_009724390.1 | 1-685 | *E. coli* BL21 | GST | nd |
| S1<sup>a</sup> | YP_009724390.1 | 16-685 | HEK293 | His | 1000 |
| S1-RBD | YP_009724390.1 | 319-541 | *E. coli* BL21 | GST | nd |
| S1-RBD | YP_009724390.1 | 1-14, 319-541 | HEK293 | His | 626 |
| S2 | YP_009724390.1 | 686-1275 | *E. coli* BL21 | GST | 798 |
| S2' | YP_009724390.1 | 817-1275 | *E. coli* BL21 | GST | 1221 |
| ORF3a | YP_009724391.1 | 1-275 | *E. coli* BL21 | GST | 287 |
| Protein E (ORF4) | YP_009724392.1 | 1-75 | *E. coli* BL21 | GST | 122 |
| Protein M (ORF5) | YP_009724393.1 | 1-222 | *E. coli* BL21 | GST | 100 |
| ORF6 | YP_009724394.1 | 1-61 | *E. coli* BL21 | GST | 100 |
| ORF7a | YP_009724395.1 | 1-121 | *E. coli* BL21 | GST | 100 |
| ORF7b | YP_009725318.1 | 1-43 | *E. coli* BL21 | GST | 100 |
| ORF8 | YP_009724396.1 | 1-121 | *E. coli* BL21 | GST | 100 |
| Protein N (ORF 9) | YP_009724397.2 | 1-419 | *E. coli* BL21 | GST | 3133 |
| N-EP3 | YP_009724397.2 | 354-400 | *E. coli* BL21 | GST | 1677 |
| ORF10 | YP_009725255.1 | 1-38 | *E. coli* BL21 | GST | 100 |
| **SARS-CoV (NC_004718.3)** |  |  |  |  |  |
| Protein N | NP_828858.1 | 1-422 | *E. coli* BL21 | GST | nd |
| **HCoV 229E (NC_002645.1)** |  |  |  |  |  |
| Protein N | NP_073556.1 | 1-389 | *E. coli* BL21 | GST | nd |
| **HCoV NL63 (NC_005831.2)** |  |  |  |  |  |
| Protein N | YP_003771.1 | 1-377 | *E. coli* BL21 | GST | nd |
| **HCoV HKU1 (NC_006577.2)** |  |  |  |  |  |
| Protein N | YP_173242.1 | 1-441 | *E. coli* BL21 | GST | nd |
| **HCoV OC43 (AY585228)** |  |  |  |  |  |
| Protein N | AAT84358.1 | 1-448 | *E. coli* BL21 | GST | Nd |

AA, amino acid; nd, not determined; <sup>a</sup>purchased from Sino Biological (Eschborn, Germany); <sup>b</sup> Mean plus three times the standard deviation in n=85 pre-pandemic (SARS-CoV-2 negative) controls for IgM/IgA/IgG antibody response; the technical minimum cut-off was 100 MFI for all antigens except S1 expressed in HEK293 cells (1000 MFI)

Since protein S1 and the sub-fragment S1-RBD were reported to be highly glycosylated (Walls et al., 2020), which might be of importance for the immunogenicity of the proteins, we included both proteins also as purified his-tagged proteins recombinantly expressed in eukaryotic HEK293 cells. S1-RBD was expressed as described previously (Stadlbauer et al., 2020) and S1 was purchased from Sino Biological (Eschborn, Germany). Indeed, bacterially expressed proteins S1 and S1-RBD showed little immunogenicity in comparison to their counterparts expressed in HEK293 cells (**Supplementary Figure S1A**). Therefore, only his-tagged S1 and S1-RBD expressed in HEK293 cells were considered for further analysis.

Antibody responses to SARS-CoV-2 protein N are speculated to potentially result from cross-reactive responses after infection with common cold Coronaviruses (ccCoVs) (Meyer et al., 2014). To assess potential cross-reactivity in the measured antibody responses, we included protein N of SARS-1 (NCBI accession no. NP_828858.1), and ccCoVs NL63 (NCBI accession no. YP_003771.1), 229E (NCBI accession no. NP_073556.1), OC43 (NCBI accession no. AAT84358.1), and HKU1 (NCBI accession no. YP_173242.1) (**Table 1**). These proteins were recombinantly expressed as GST-tagged fusion proteins in *E. coli* BL21 as described above.

### Multiplex Serology

The GST-tagged fusion proteins were affinity-purified on glutathione-casein coated fluorescently labelled polystyrene beads (Luminex Corp., Austin, TX, USA), whereas purified his-tagged proteins were directly cross-linked to the bead surface, as described previously (Butt et al., 2016; Waterboer et al., 2005).

Mixing the bead sets loaded with distinct antigens allowed a high-throughput simultaneous analysis of several antigens per serum. For all secondary antibodies, sera were pre-incubated (1:50 dilution) in a buffer containing polyvinylalcohol, polyvinylpyrrolidon, casein, and protein lysate of *E. coli* over-expressing GST-tag to suppress unspecific binding of antibodies to the glutathione-coated beads and residual *E. coli* proteins. For simultaneous detection of IgM/IgA/IgG, sera were additionally pre-incubated with 2.5% Super ChemiBlock™ Heterophile Blocking Agent (CBS-K, Chemicon, Temecula, CA, USA) (Waterboer et al., 2006), and for IgM with 1:10 diluted Rf-absorbens (Linaris, Mannheim, Germany). After the pre-incubation step, sera were mixed (1:1) and incubated with the antigen-loaded bead mixture. Bound serum antibodies were labelled separately with biotinylated secondary antibodies (goat anti-human IgM/IgA/IgG, anti-human IgM, anti-human IgG, or anti-human IgA; Jackson ImmunoResearch, Westgrove, PA, USA) and subsequently incubated with Streptavidin-R-Phycoerythrin (MossBio, Pasadena, MD, USA). Simultaneous detection of IgM/IgA/IgG is reported unless otherwise specified. A Luminex 200 Analyzer (Luminex Corp., Austin, TX, USA) was used to distinguish the bead sets and their respective antigens, and to quantify the amount of bound serum antibody. The level of antibody response is given as median fluorescence intensity (MFI) of at least 100 beads per type measured. Background values against the GST-tag, as well as the bead-surface and secondary reagents were subtracted to generate net MFI values.

Antigen-specific cut-offs for sero-positivity (**Table 1**) were defined as the mean plus three times the standard deviation among the analyzed population sampled before the Covid-19 pandemic (pre-pandemic controls). The technical minimum cut-off was 100 MFI, except for Protein S1 expressed in HEK293 cells (1000 MFI) due to observed variations in background values.

### SARS-CoV-2 IgG ELISA

For n=124 of the total of n=241 serum samples analyzed in multiplex serology, we performed a concurrent SARS-CoV-2 IgG enzyme-linked immunosorbent assay (ELISA) specific for antibodies directed against the S1 domain of the spike protein (Euroimmun, Lübeck, Germany). The samples were analyzed on a Euroimmun Analyzer I instrument according to the manufacturer’s instructions. Briefly, serum was applied in a 1:101 dilution on the provided antigen-coated ELISA plates, and subsequently incubated with a peroxidase-linked anti-human IgG secondary antibody. The amount of bound serum antibody was measured as the optical density (OD) at 450 nm after incubation with substrate solution. The ratio of the obtained OD per sample and the extinction of a calibrator sample were calculated. Following the manufacturer’s protocol, a ratio < 0.8 was considered sero-negative, a ratio ≥ 1.1 was considered sero-positive, and ratios between 0.8 and 1.0 considered borderline.

### Microscopy-based immunofluorescence detection of IgG antibodies to SARS-CoV-2

Microscopy-based immunofluorescence detection of IgG antibodies to SARS-CoV-2 antibodies was performed using a newly established semi-automated, semi quantitative approach, as described previously (Pape et al., 2020), for n=38 serum samples of hospitalized Covid-19 patients. In contrast to the ELISA measurements this approach is not specific for a single viral protein, but can detect antibodies against all viral proteins expressed in the infected cell context. Briefly, fixed and permeabilized SARS-CoV-2 infected VeroE6 cells in 96-well plates were incubated with patient serum or control serum, respectively. Bound serum IgG antibodies to SARS-CoV-2 were detected using goat anti-human IgG-AlexaFluor 488 (Invitogen, Thermofisher Scientific) and evaluated by fluorescence microscopy. Concomitant immunostaining against dsRNA was performed in order to differentiate between virus-expressing cells (dsRNA positive) and non-infected cells (dsRNA negative) in the same specimen. Semi-automated microscopy and image data analysis was performed as described in detail in (Pape et al., 2020). For each sample, a score was calculated based on the ratio of immunostaining intensity determined for cells infected with SARS-CoV-2 to that of the non-infected cells in the same specimen. A threshold-value of 1.30 for sero-positivity was defined in (Pape et al., 2020) based on receiver operating characteristic (ROC) analysis to achieve optimum specificity. Scores < 1.27 were considered sero-negative, and those between 1.27 and 1.30 as borderline to allow for better sensitivity of the assay (Pape et al., 2020).

### Statistical Analysis

Continuous antibody responses to SARS-CoV-2 proteins were compared between pre-pandemic controls and hospitalized Covid-19 patients using the Wilcoxon rank sum test. A p-value < 0.05 was considered statistically significant. Agreement between SARS-CoV-2 multiplex serology and commercial SARS-CoV-2 ELISA (Euroimmun, Lübeck, Germany) was assessed using Cohen’s kappa and the respective 95% confidence interval (CI).

All graphical and descriptive representations were performed using SAS 9.4 (SAS Institute, Cary, NC, USA) or GraphPad Prism 8 (Graphpad Software, Inc., San Diego, CA, USA).

All summary data generated or analyzed during this study are included in this published article and its supplementary material file. The primary data are available from the corresponding author on reasonable request.

## RESULTS

### Antibody responses to SARS-CoV-2 proteins in pre-pandemic controls and hospitalized Covid-19 patients

Antibody responses to all proteins included in SARS-CoV-2 multiplex serology were compared between n=85 pre-pandemic controls and n=48 hospitalized Covid-19 patients based on a single serum sample drawn at the latest time point after symptom onset (range 2 to 43 days) (**Figure 1**).

**Figure 1:**
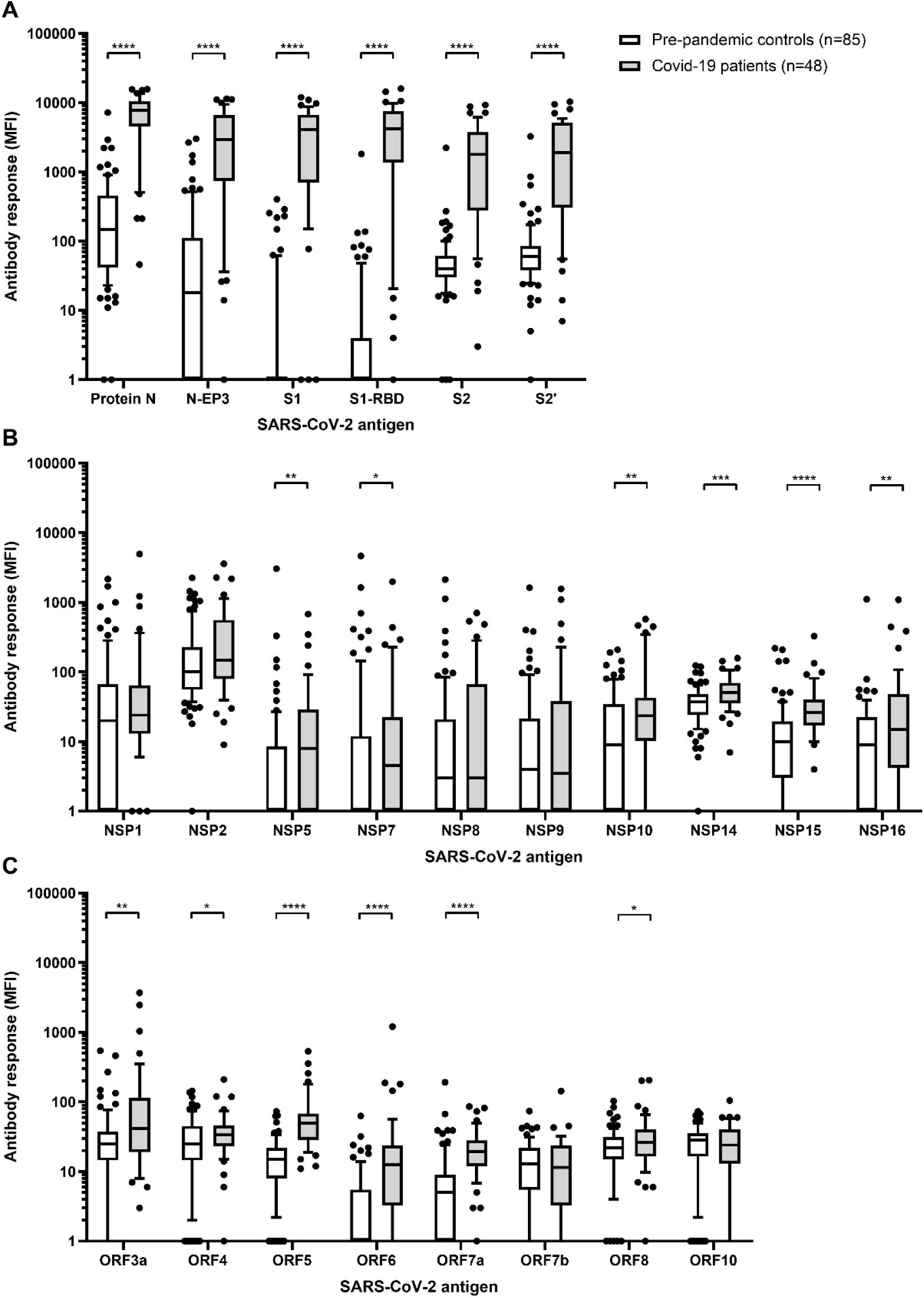
Antibody responses to SARS-CoV-2 proteins in n=85 pre-pandemic controls and n=48 hospitalized Covid-19 patients. A) Antibody responses (MFI) to proteins N, S1 and S2, as well as their respective fragments; B) SARS-CoV-2 non-structural proteins (NSP); C) other SARS-CoV-2 open reading frames (ORF). If multiple samples per individual were available only the sample with the latest time point after blood draw was considered for analysis. Boxes represent the 25^th^ to 75^th^ and whiskers the 10^th^ to 90^th^ percentiles, respectively. Wilcoxon rank sum test was applied to assess statistically significant differences in antibody responses (MFI) to SARS-CoV-2 proteins between pre-pandemic controls and hospitalized Covid-19 patients; *p-value < 0.05, **p-value < 0.01, ***p-value < 0.001, ****p-value < 0.0001.

Among controls, protein N exerted the highest antibody response of all proteins analyzed (median: 149 MFI, range 1 – 7195 MFI) (**Figure 1A**). This was also reflected in the comparably high cut-off of 3133 MFI determined as the mean plus three times the standard deviation among pre-pandemic controls (**Table 1**). For all other proteins, the median antibody response ranged between 1 and 101 MFI, and the cut-off values were much lower.

We further assessed whether the observed high responses to protein N result from cross-reactive antibody responses potentially originating from previous infections with SARS-CoV-1 and endemic ccCoVs NL63, HKU1, 229E and OC43. Antibody responses to proteins N of OC43, HKU1, NL63, and 229E were abundantly high in pre-pandemic controls (> 1000 MFI) and consequently there was little correlation with antibody responses to protein N of SARS-CoV-2 (**Supplementary Figure S2B and C)**. However, antibody responses to the N proteins of SARS-CoV-1 and SARS-CoV-2 were correlated (R^2^ = 0.55) (**Supplementary Figure S2A**).

Proteins with the highest levels of immunogenicity in Covid-19 patients were proteins N, S1, and S2 (median: 7825, 4098, and 1789 MFI, respectively) as well as their respective sub-fragments N-EP3, S1-RBD, and S2’ (median: 2943, 4228, and 1030 MFI, respectively) (**Figure 1A**). Antibody responses to these proteins were all significantly higher among Covid-19 patients compared to pre-pandemic controls (all p-values < 0.0001, **Figure 1A**). For all other proteins included in the multiplex serology assay, except NSP1, NSP2, NSP8, NSP9, ORF7a, and ORF10, antibody responses were also significantly higher among cases than controls (all p-values < 0.05). However, overall antibody responses to these proteins were comparably low among cases with medians ranging between 4 and 147 MFI, and mostly below the determined cut-off values (**Table 1**).

Since the Covid-19 patients in our cohort were sampled at various time points after symptom onset, we analyzed the available longitudinal samples for sero-conversion (**Figure 2** and **Supplementary Figure S3**). Indeed, for the most immunogenic proteins N, N-EP3, S1, S1-RBD, S2, and S2’ the majority of hospitalized Covid-19 patients sero-converted at the latest two weeks after symptom onset (**Figure 2**). For all other proteins, sero-conversion was a rare event and was only observed for individual patients at distinct time-points after symptom onset (**Supplementary Figure S3**).

**Figure 2:**
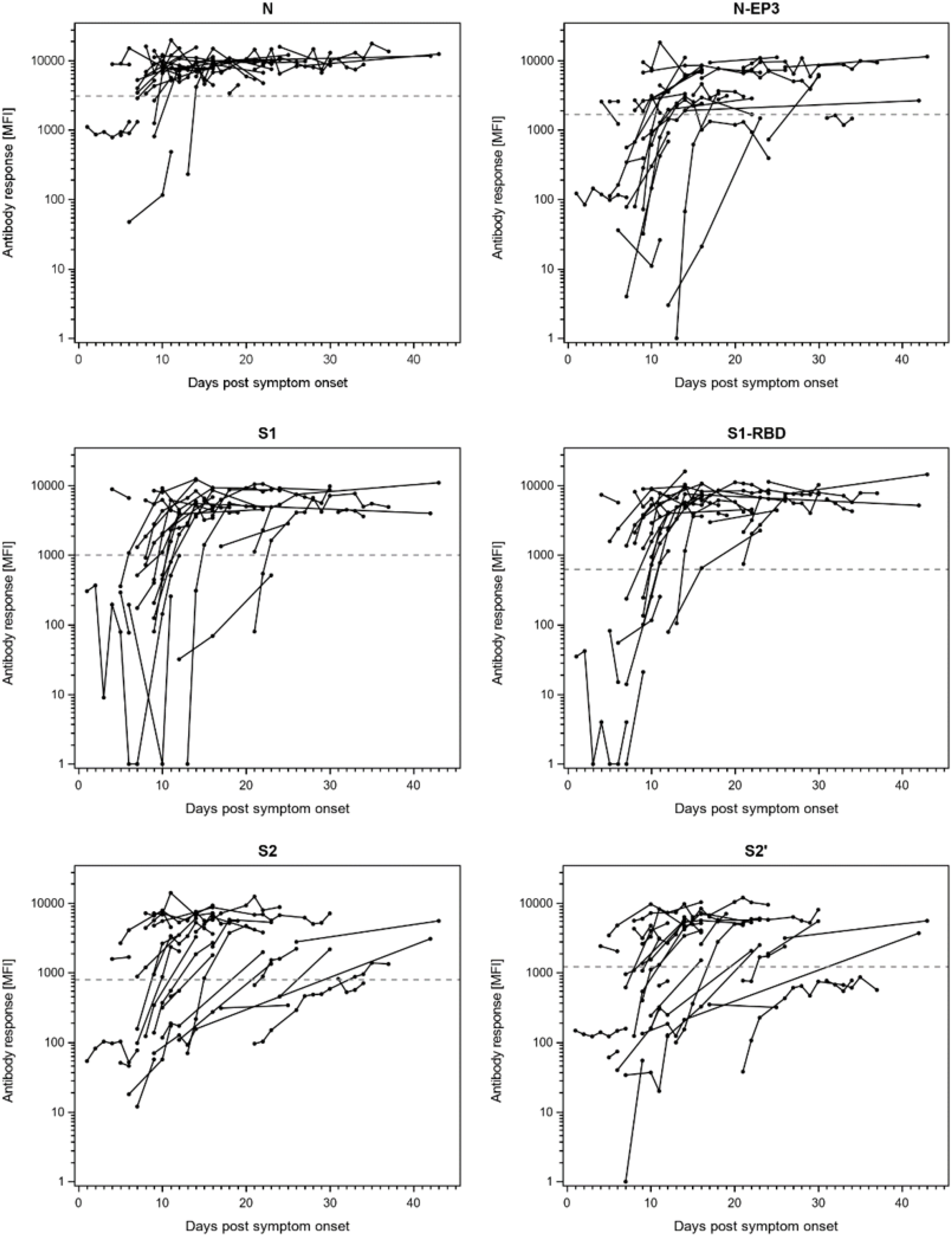
Antibody responses to SARS-CoV-2 proteins N, S1, S2, and their respective sub-fragments (N-EP3, S1-RBD, and S2’) in longitudinal samples of n=32 hospitalized Covid-19 patients. The antibody response (MFI) is plotted against days post symptom onset. The dashed lines indicate the antigen-specific cut-offs defined as mean plus three times the standard deviation in n=85 pre-pandemic controls.

### Assessment of sensitivity and specificity of the newly developed SARS-CoV-2 multiplex serology assay

Based on the specific timing of sero-conversion to SARS-CoV-2 proteins described above, we assessed sensitivity of the newly developed multiplex serology assay separately for hospitalized Covid-19 patients, who provided their latest blood sample ≤14 days (n=23) or >14 days (n=25) after symptom onset (**Table 2**). Among patients with a blood sample drawn >14 days after symptom onset, the highest sero-prevalence was achieved with proteins N and S1-RBD (both 100%, 95% CI: 86%-100%), followed by protein S1 (96%, 95% CI: 80%-99%), proteins S2 and N-EP3 (both 80%, 95% CI: 59%-93%), and protein fragment S2’ (76%, 95% CI: 55%-91%). Sero-prevalence for the other SARS-CoV-2 proteins was low and reached at most 16% (n=4 sero-positive individuals) among hospitalized patients with a blood sample drawn >14 days after symptom onset.

**Table 2:** Sero-prevalence of SARS-CoV-2 proteins in pre-pandemic controls, hospitalized and non-hospitalized Covid-19 patients.

| Antigen | N (%) sero-positive |  |  |  |
| --- | --- | --- | --- | --- |
|  | Controls<br>(n=85) | Patients, non-hospitalized<br>(n=15)<sup>a,b</sup> | Hospitalized patients,<br>≤14 days post<br>symptom onset (n=23)<sup>b</sup> | Hospitalized patients,<br>>14 days post<br>symptom onset (n=25)<sup>b</sup> |
| Protein N | 1 (1) | 1 (7) | 13 (57) | 25 (100) |
| N-EP3 | 3 (4) | 0 (0) | 9 (39) | 20 (80) |
| S1 | 0 (0) | 0 (0) | 11 (48) | 24 (96) |
| S1-RBD | 1 (1) | 3 (20) | 13 (57) | 25 (100) |
| S2 | 1 (1) | 1 (7) | 10 (43) | 20 (80) |
| S2' | 1 (1) | 0 (0) | 9 (39) | 19 (76) |
| Protein N + S1-RBD | 0 (0) | 1 (7) | 11 (48) | 25 (100) |
| NSP1 | 2 (2) | 1 (7) | 1 (4) | 1 (4) |
| NSP2 | 2 (2) | 0 (0) | 2 (9) | 1 (4) |
| NSP5 | 1 (1) | 0 (0) | 0 (0) | 0 (0) |
| NSP7 | 1 (1) | 0 (0) | 0 (0) | 1 (4) |
| NSP8 | 2 (2) | 0 (0) | 0 (0) | 0 (0) |
| NSP9 | 1 (1) | 0 (0) | 2 (9) | 0 (0) |
| NSP10 | 2 (2) | 0 (0) | 4 (17) | 4 (16) |
| NSP14 | 2 (2) | 0 (0) | 0 (0) | 4 (16) |
| NSP15 | 4 (5) | 1 (7) | 1 (4) | 0 (0) |
| NSP16 | 1 (1) | 0 (0) | 2 (9) | 0 (0) |
| ORF3a | 2 (2) | 0 (0) | 2 (9) | 3 (12) |
| ORF4 | 2 (2) | 1 (7) | 1 (4) | 0 (0) |
| ORF5 | 0 (0) | 0 (0) | 4 (17) | 2 (8) |
| ORF6 | 0 (0) | 0 (0) | 2 (9) | 2 (8) |
| ORF7a | 1 (1) | 0 (0) | 0 (0) | 0 (0) |
| ORF7b | 0 (0) | 0 (0) | 1 (4) | 0 (0) |
| ORF8 | 1 (1) | 0 (0) | 1 (4) | 1 (4) |
| ORF10 | 0 (0) | 0 (0) | 1 (4) | 1 (4) |

<sup>a</sup>all sampled within two weeks after symptom onset; <sup>b</sup>one sample per individual drawn at the latest time point after symptom onset.

Antibody responses to S1 and S1-RBD strongly correlated with each other (R^2^=0.93) (**Supplementary Figure S1B**). Since S1-RBD showed a slightly higher prevalence in hospitalized patients with blood drawn ≤14 days after symptom onset (57%, 95% CI: 34%-77%) than S1 (48%, 95% CI: 27%-69%), we considered S1-RBD as parameter for a final algorithm defining SARS-CoV-2 sero-positivity (**Table 2**).

Protein N-EP3 showed substantially lower sensitivity than the full length protein N (80% versus 100%, respectively); and proteins S2 and S2’ also only achieved sensitivities of up to 80% in hospitalized patients more than two weeks after symptom onset. Consequently, proteins N-EP3, S2, and S2’ do not further contribute to an increased sensitivity in addition to Protein N and S1-RBD. Defining SARS-CoV-2 overall sero-positivity as being dual-positive to Protein N and S1-RBD resulted in 100% sensitivity (95% CI: 86%-100%) in detecting hospitalized Covid-19 patients >14 days after symptom onset. Among hospitalized patients with a shorter time between symptom onset and blood draw, the sensitivity of our assay was 48% (95% CI: 27%-69%). Interestingly, among non-hospitalized patients (n=15), all sampled within 14 days after symptom onset, only one individual (7%) was considered sero-positive with SARS-CoV-2 multiplex serology.

Among pre-pandemic controls, only one individual each was sero-positive to N or S1-RBD, respectively. However, none of the pre-pandemic controls was dual-positive and thus considered SARS-CoV-2 sero-positive, indicating 100% (95% CI: 96%-100%) specificity of our newly developed assay.

### Comparison of SARS-CoV-2 multiplex serology to a commercially available ELISA and microscopy-based immunofluorescence

We generated concurrent data from a widely used commercial SARS-CoV-2 IgG ELISA (Euroimmun, Lübeck, Germany) for n=124 serum samples (n=30 pre-pandemic controls, n=88 serum samples from n=25 hospitalized Covid-19 patients, and n=6 serum samples from n=5 non-hospitalized Covid-19 patients). Two of the analyzed serum samples were classified as borderline in ELISA. These samples consisted of one pre-pandemic control, who was considered negative with SARS-CoV-2 multiplex serology (Protein N + S1-RBD sero-negative), and one Covid-19 patient, who was defined positive with SARS-CoV-2 multiplex serology.

The ELISA definition for being sero-negative or -positive for SARS-CoV-2 was in substantial agreement (Cohen’s kappa = 0.68; 95% CI: 0.56-0.80) with being dual-positive to protein N and S1-RBD in multiplex serology (**Table 3**). All ELISA sero-positives (n=49) were also sero-positive in multiplex serology. However, n=20 of the n=73 ELISA sero-negatives were defined positive in multiplex serology. These 20 samples consisted of n=19 samples from hospitalized Covid-19 patients and n=1 sample from a non-hospitalized Covid-19 patient with mild symptoms.

**Table 3:** Comparison of SARS-CoV-2 sero-positivity in multiplex serology with commercially available ELISA and microscopy-based immunofluorescence detection of antibodies to SARS-CoV-2.

|  | N (%) | ELISA (IgG)* |  |  | Microscopy-based immunofluorescence (IgG) |  |  |
| --- | --- | --- | --- | --- | --- | --- | --- |
|  |  | Negative | Positive | Total | Negative | Positive | Total |
| **SARS-CoV-2 multiplex serology, Protein N + S1-RBD** | **Negative** | 53 (44) | 0 (0) | 53 (44) | 10 (27) | 1 (3) | 10 (30) |
|  | **Positive** | 20 (16) | 49 (40) | 69 (56) | 0 (0) | 26 (70) | 26 (70) |
|  | **Total** | 73 (60) | 49 (40) | 122 (100) | 9 (27) | 27 (73) | 37 (100) |

\*Euroimmun (Lübeck, Germany)

For n=38 samples of hospitalized Covid-19 patients, we were able to compare our results to microscopy-based immunofluorescence detection of IgG antibodies to SARS-CoV-2. Of these, n=1 sample was considered borderline in the microscopy-based assay but sero-negative in multiplex serology. For the remaining n=37 samples, the microscopy-based assay and multiplex serology were in almost perfect agreement (Cohen’s kappa = 0.93; 95% CI: 0.81-1.00). Only one sample had a discordant result between the two assays, and was considered sero-positive in the microscopy-based assay but sero-negative in multiplex serology (**Table 3**).

### Sero-positivity to SARS-CoV-2 proteins other than N and S and severity of Covid-19 disease

In addition to whether patients were hospitalized, further information was available on oxygenation requirements and disease outcome of hospitalized patients. In an attempt to identify serological markers for disease severity, we included almost the entire SARS-CoV-2 proteome in our multiplex serology. Due to the small sample size, it was impossible to generate robust age-and sex-adjusted regression models, i.e. we were not able to adequately control for differences in patient characteristics. Therefore, we present a purely descriptive analysis (**Table 4**). Overall, sero-positivity to SARS-CoV-2 proteins other than N, N-EP3, S1, S1-RBD, S2, and S2’ was a rare event and observed in n=10 hospitalized patients (20%) and in none of the non-hospitalized patients. Although all patients sero-positive to ≥2 SARS-CoV-2 proteins other than N and S or their sub-fragments were hospitalized (n=10), the majority of them had a more favorable course of disease. N=9 of these 10 patients were among the n=45 patients that needed oxygenation during their hospital stay, however, 8 out of these 9 needed only oxygenation through a nasal cannula, in contrast to High-Flow Nasal Oxygen (HFNO) or invasive oxygenation. Of note, the single patient who did not need any oxygenation was sero-positive to the maximum number of proteins observed in this study (n=5). Furthermore, none of the hospitalized patients that were sero-positive to at least two of the additional proteins died due to Covid-19.

**Table 4:** Age, sex, and sero-positivity to SARS-CoV-2 proteins other than protein N and S and their sub-domains and severity of disease in studied Covid-19 patients.

|  | Hospitalized |  | Oxygenation<sup>b</sup> |  | Type of oxygenation<sup>c</sup> |  | Death<sup>b</sup> |  |
| --- | --- | --- | --- | --- | --- | --- | --- | --- |
|  | No<br>(n=15) | Yes<br>(n=48) | No<br>(n=5) | Yes<br>(n=43) | Nasal<br>cannula<br>(n=23) | HFNO/<br>Invasive<br>(n=20) | No<br>(n=41) | Yes<br>(n=7) |
| Age [years], median (range) | 54 (27-70) | 62 (23-85) | 50 (23-61) | 63 (36-85) | 55 (36-75) | 69 (42-85) | 59 (23-79) | 78 (72-85) |
| Sex, n (%) female | 9 (60%) | 16 (33%) | 4 (80%) | 12 (28%) | 8 (35%) | 4 (20%) | 15 (36%) | 1 (14%) |
| Number of sero-positive<br>proteins<sup>a</sup>, n (%) |  |  |  |  |  |  |  |  |
| 0 | 12 (80) | 22 (46) | 3 (60) | 19 (44) | 8 (35) | 11 (55) | 17 (41) | 5 (71) |
| 1 | 3 (20) | 16 (34) | 1 (20) | 15 (35) | 7 (30) | 8 (40) | 14 (34) | 2 (29) |
| 2 | 0 (0) | 5 (10) | 0 (0) | 5 (12) | 4 (17) | 1 (5) | 5 (12) | 0 (0) |
| 3 | 0 (0) | 4 (8) | 0 (0) | 4 (9) | 4 (17) | 0 (0) | 4 (10) | 0 (0) |
| 4 | 0 (0) | 0 (0) | 0 (0) | 0 (0) | 0 (0) | 0 (0) | 0 (0) | 0 (0) |
| 5 | 0 (0) | 1 (2) | 1 (20) | 0 (0) | 0 (0) | 0 (0) | 1 (2) | 0 (0) |

HFNO, High-Flow Nasal Oxygen; <sup>a</sup>among all proteins except N, N-EP3, S1, S1-RBD, S2, and S2'; <sup>b</sup>among hospitalized patients; <sup>c</sup>among those patients that needed oxygenation

### Separate detection of immunoglobulin M, A, and G antibody responses in Covid-19 patients with longitudinal samples

Besides measuring Ig classes M, A, and G simultaneously, we also assessed the antibody response to SARS-CoV-2 proteins separately by Ig class (**Supplementary Figure S4**). We here exemplarily show IgM, IgA, and IgG responses to proteins N and S1-RBD for three hospitalized Covid-19 patients with multiple follow-up samples between days 9 and 29 (patient 1), 8 and 24 (patient 2), and 21 and 37 (patient 3) post symptom onset. The antibody responses by Ig class appeared distinct for the two proteins. In case of protein N, the IgM, IgA, and IgG response remained fairly stable over the course of follow-up for all three patients. For S1-RBD, however, the IgM and IgA responses decreased by about 50% over the course of follow-up for patients 1 and 2, whereas IgG responses remained stable or even increased. For patient 3, IgM and IgA remained stable, whereas IgG level increased from days 21 to 37 post symptom onset.

## DISCUSSION

We here present the development of a multiplex serology assay for detection of antibody responses to SARS-CoV-2. Defining dual sero-positivity to the nucleocapsid protein N and the RBD domain of spike protein S for classification as a positive result obtained 100% sensitivity and specificity in hospitalized Covid-19 patients and pre-pandemic controls. The assay was furthermore in excellent agreement with other serological SARS-CoV-2 assays. Inclusion of additional SARS-CoV-2 proteins in the antigen panel as well as assessment of different Ig classes allowed for further explorative analyses with progression of disease and course of antibody response.

The current SARS-CoV-2 pandemic leads to an urgent need for high-throughput approaches to assess SARS-CoV-2 epidemiology. We developed a multiplex serology assay based on fluorescent-bead based technology that includes the well-described SARS-CoV-2 immunogenic proteins N and S1 as well as their respective subdomains N-EP3 and S1-RBD (Grifoni et al., 2020; Post et al., 2020; Stadlbauer et al., 2020). We found that dual sero-positivity to proteins N and S1-RBD resulted in 100% sensitivity and 100% specificity when applied in hospitalized Covid-19 patients sampled >14 days after symptom onset and age- and sex-matched pre-pandemic controls, respectively. Classification of SARS-CoV-2 sero-positivity separately by protein N or S1-RBD would have resulted in false-positive classifications of each one pre-pandemic control and consequently a lower specificity, highlighting the benefit of simultaneously assessing multiple antigens. This is further supported by comparison of SARS-CoV-2 multiplex serology with commercially available ELISA, detecting antibody responses to protein S1 only. Comparison of the S1-RBD protein applied in multiplex serology to ELISA results in substantial agreement (Cohen’s kappa = 0.65; 95% CI: 0.53-0.77) between the two assays; very similar to the comparison based on N and S1-RBD dual sero-positivity, i.e. the definition of overall SARS-CoV-2 sero-positivity in multiplex serology. However, one pre-pandemic control being defined as borderline in ELISA was defined sero-positive with the S1-RBD measurement in multiplex serology (data not shown). With the dual-classification for sero-positivity to proteins N and S1-RBD, this pre-pandemic control was correctly identified as sero-negative.

With respect to specificity of the assay, the potential of cross-reactive antibody responses resulting from previous infections with other CoVs including SARS-CoV-1 and endemic ccCoVs NL63, HKU1, 229E and OC43, should be acknowledged (Becker et al., 2020; Meyer et al., 2014). We observed the highest antibody responses among pre-pandemic controls to the nucleocapsid protein N, which exerts a strong potential for cross-reactive antibody responses due to high sequence similarity between related CoVs (Meyer et al., 2014). We therefore expressed the respective N proteins of SARS-CoV-1, OC43, HKU1, NL63, and 229E to address this issue. Antibody responses to proteins N of OC43, HKU1, NL63, and 229E were abundantly high (above 1000 MFI for the majority of pre-pandemic controls) and consequently there was little correlation with antibody responses to protein N of SARS-CoV-2, which is in concordance with one previously published study (Becker et al., 2020). We can therefore not exclude that high antibody responses to SARS-CoV-2 protein N among controls may result from a previous infection with other CoVs, the most likely candidates being the ccCoVs (Edridge et al., 2020). However, our data also shows that not all high antibody responses to protein N of other CoVs necessarily induce cross-reactive responses to SARS-CoV-2. Antibody responses to the N proteins of SARS-CoV-1 and SARS-CoV-2 correlated (R^2^ = 0.55) but we do not expect any previous SARS-CoV-1 patients among our control group, since these were sampled before the SARS-CoV-1 epidemic in 2002/2003. Moreover, the SARS-CoV-1 pandemic was mostly localized in South-East Asia and our samples (controls and cases) were sampled in Germany. Thus, antibody responses to protein N most likely do not originate from previous SARS-CoV-1 infections in our population, however, they should be considered when analyzing studies from the respective time window and geographical regions. Based on our preliminary observations presented here, it will be important to include also the homologous spike proteins into the multiplex assay to more comprehensively assess potential cross-reactive antibody responses.

The newly developed SARS-CoV-2 multiplex serology not only provided the potential to increase specificity of SARS-CoV-2 serology but also showed a high sensitivity in detection of SARS-CoV-2 infected individuals. In comparison to above-mentioned commercially available ELISA, we detected n=20 (16%) discordant samples that were sero-negative in ELISA but classified sero-positive in SARS-CoV-2 multiplex serology. Of note, in samples that were part of a longitudinal series in patients (for 11 of these 20 samples), multiplex serology was able to identify Covid-19 patients as sero-positive up to 8 days earlier post symptom onset than the ELISA, indicating a higher sensitivity of SARS-CoV-2 multiplex serology. This might be in part due to the choice of secondary antibody. The commercially available ELISA used in this study detects IgG antibody responses only, whereas our secondary antibody for classification of SARS-CoV-2 sero-positivity detects IgM/IgA/IgG simultaneously. We performed a separate analysis with SARS-CoV-2 multiplex serology focusing on IgG and found both tests also to be in substantial agreement (Cohen’s kappa = 0.77; 95% CI: 0.66-0.88). However, for the IgG measurement, only n=14 (versus n=20 with simultaneous IgM/IgA/IgG detection) of the hospitalized Covid-19 patients were defined ELISA sero-negative but multiplex serology sero-positive. Similarly, the microscopy-based immunofluorescence detection of IgG antibodies to SARS-CoV-2 in comparison to the IgG measurement in multiplex serology showed two hospitalized Covid-19 patients being sero-negative in IgG multiplex serology but sero-positive in immunofluorescence, as opposed to only one discordant patient with the IgM/IgA/IgG measurement. Thus, application of a secondary antibody that detects multiple Ig isotypes simultaneously potentially allows for detection of individuals that had undergone SARS-CoV-2 infection that otherwise might have been missed, as indicated in the literature (Hu et al., 2020).

The application of a multiplex technique to detect antibody responses to SARS-CoV-2 has further advantages including the assessment of additional SARS-CoV-2 proteins that might not be relevant for classification of sero-positivity but are potentially associated with disease course and/or progression, hence our reference to “serolomics”, a term that has been initially coined for Human Papillomavirus multiplex serology (Dillner, 2005). We here performed an explorative analysis and described the frequency of individuals with sero-positivity to multiple SARS-CoV-2 proteins other than the nucleocapsid or spike proteins, including non-structural proteins and other ORFs. Overall, n=10 of hospitalized patients and none of the non-hospitalized patients were sero-positive to ≥2 SARS-CoV-2 proteins other than proteins N and S or their sub-fragments. Although sero-positivity to ≥2 SARS-CoV-2 proteins other than proteins N and S or their sub-fragments occurred only among a proportion of hospitalized patients, the majority of them had an overall more favorable course of disease including less invasive need for oxygenation (n=9). Moreover, none of the patients with antibody responses to multiple additional SARS-CoV-2 proteins died of Covid-19. Of note, the different patient groups in this analysis differed also in age and sex (**Table 4**) and due to the small sample size we were not able to appropriately control for patient characteristics in age- and sex-adjusted regression models. However, these findings raised our specific interest and should be analyzed more deeply in larger patient cohorts. A potential underlying biological mechanism for the above-described observation of a more favorable course of disease with a more pronounced antibody response to the SARS-CoV-2 proteome might be correlating B- and T-cell responses contributing to viral clearance (Ferretti et al., 2020).

Our study has several strengths and limitations. One major limitation is the fact that we only detected one out of 15 non-hospitalized Covid-19 patients as sero-positive. However, it should be noted that the non-hospitalized patients included in our study were all sampled within two weeks after symptom onset. In concordance with the literature (Okba et al., 2020), we observed that among hospitalized patients only after 14 days post symptom onset maximum sensitivity of the assay can be achieved. Moreover, non-hospitalized patients were of younger age and had a higher proportion of females than our control population, which might have affected the level of antibody response. A larger pre-pandemic control sample set with a wider age distribution is needed to address this question. The sensitivity among SARS-CoV-2 infected individuals experiencing mild or no symptoms also in relation to the time-point of infection needs to be further elucidated, especially with respect to future applications of the assay in large population-based cohorts. A major strength of our newly developed SARS-CoV-2 serological assay is its multiplex approach, allowing for the assessment of up to 100 antigens in parallel. It does not only permit inclusion of multiple SARS-CoV-2 proteins but also autoantigens or antigens from pathogens that might induce antibody responses due to (re-)activation or co-infection during the course of Covid-19 disease, as it was previously described for Herpesviruses (Amorim Dos Santos et al., 2020; Elsaie et al., 2020; Xu et al., 2020). We furthermore demonstrated the possibility to quantify longitudinally the humoral anti-SARS-CoV-2 responses for different Ig isotypes. Analysis of samples from larger cohorts of Covid-19 patients with long-term follow-up will provide information on the natural development and stability of anti-SARS-CoV-2 responses for all Ig isotype classes.

In conclusion, we here present the development of a multiplex serology assay that covers a substantial proportion of the SARS-CoV-2 proteome. Considering the achieved high sensitivity and specificity of the assay and its high-throughput applicability in large sero-epidemiological studies this assay might be suitable for efficient analysis of SARS-CoV-2 epidemiology in population-based studies. The option of hypothesis-driven inclusion of additional antigens into the panel as well as separate detection of different Ig isotypes will furthermore allow the assessment of additional research questions.

