## Supplementary Material for "From multiplex serology to serolomics – a novel approach to the antibody response against the SARS-CoV-2 proteome"



**Funding:**

This work was generously supported by the Dieter Morszeck Stiftung. Data and sample collection for the ESTHER study were supported by a grant from the Baden-Württemberg State Ministry of Science, Research and Arts.

### SUPPLEMENTARY MATERIAL

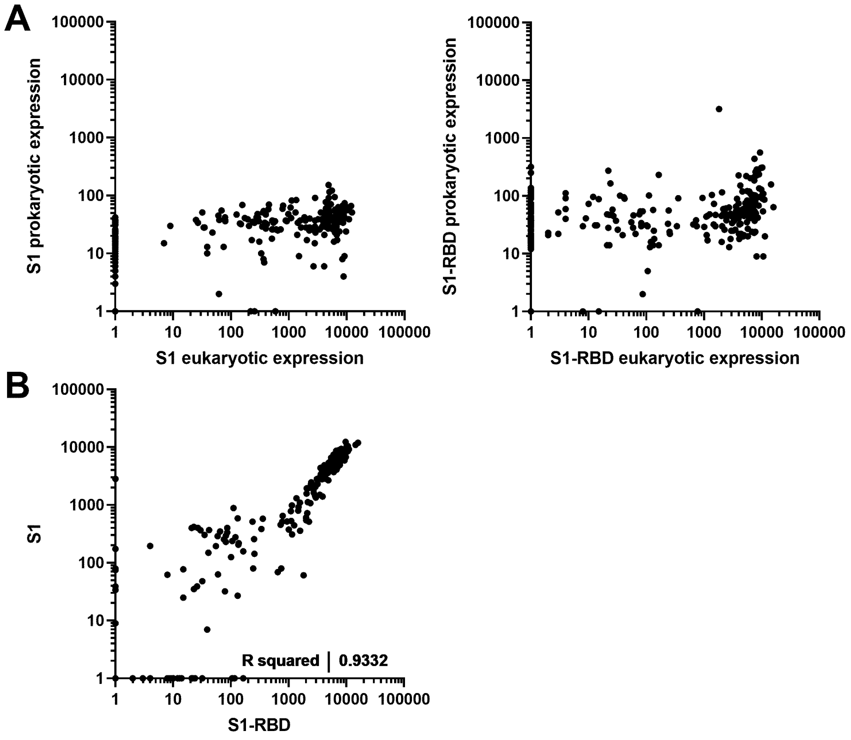


**Supplementary figure S1: Antibody responses to SARS-CoV-2 proteins S1 and S1-RBD in n=174 serum samples of Covid-19 patients and n=85 serum samples of pre-pandemic controls.** A) Comparison of antibody responses (MFI) to S1 and S1-RBD proteins expressed in HEK293 (eukaryotic expression) to those expressed in *E. coli* BL21 (prokaryotic expression); B) Correlation of antibody responses (MFI) to eukaryotically expressed S1 and S1-RBD; the Pearson correlation coefficient (R squared) is given as a measure for correlation.

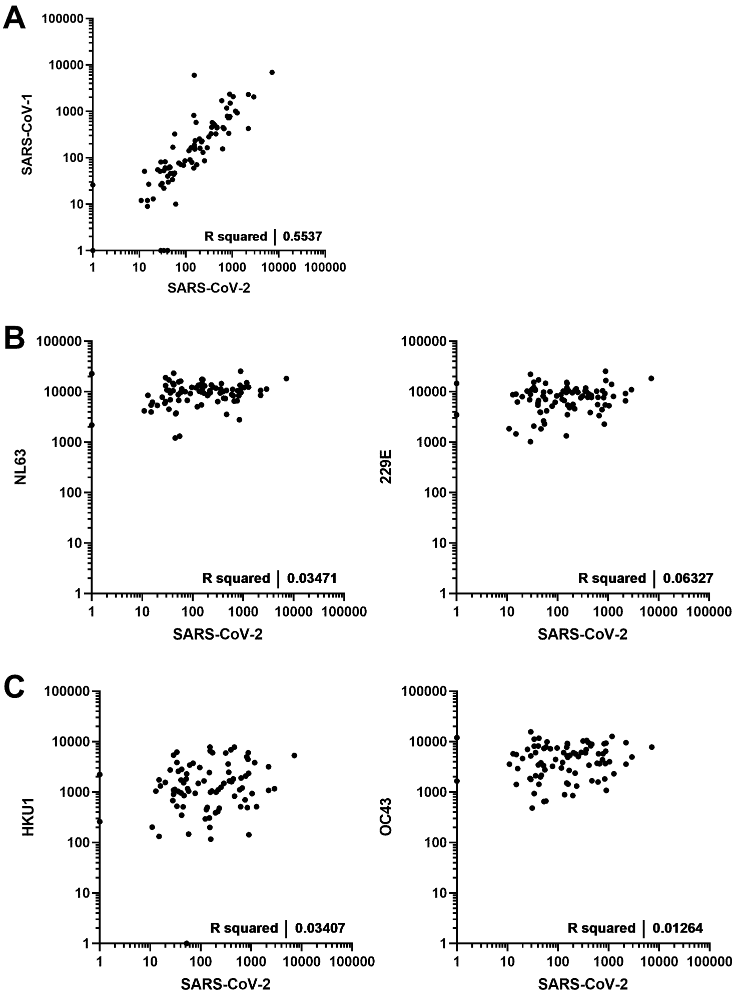


**Supplementary figure S2: Correlation of antibody responses to protein N of SARS-CoV-2 to those of other CoVs.** Antibody responses (MFI) to protein N of SARS-CoV-2 are compared to those of SARS-CoV-1 and ccCoVs: A) SARS-CoV-1, B) α-CoVs NL63 and 229E, and c) β-CoVs OC43 and HKU1. Blood samples were drawn prior to the Covid-19 pandemic (n=85). The Pearson correlation coefficient (R squared) is given as a measure for correlation.

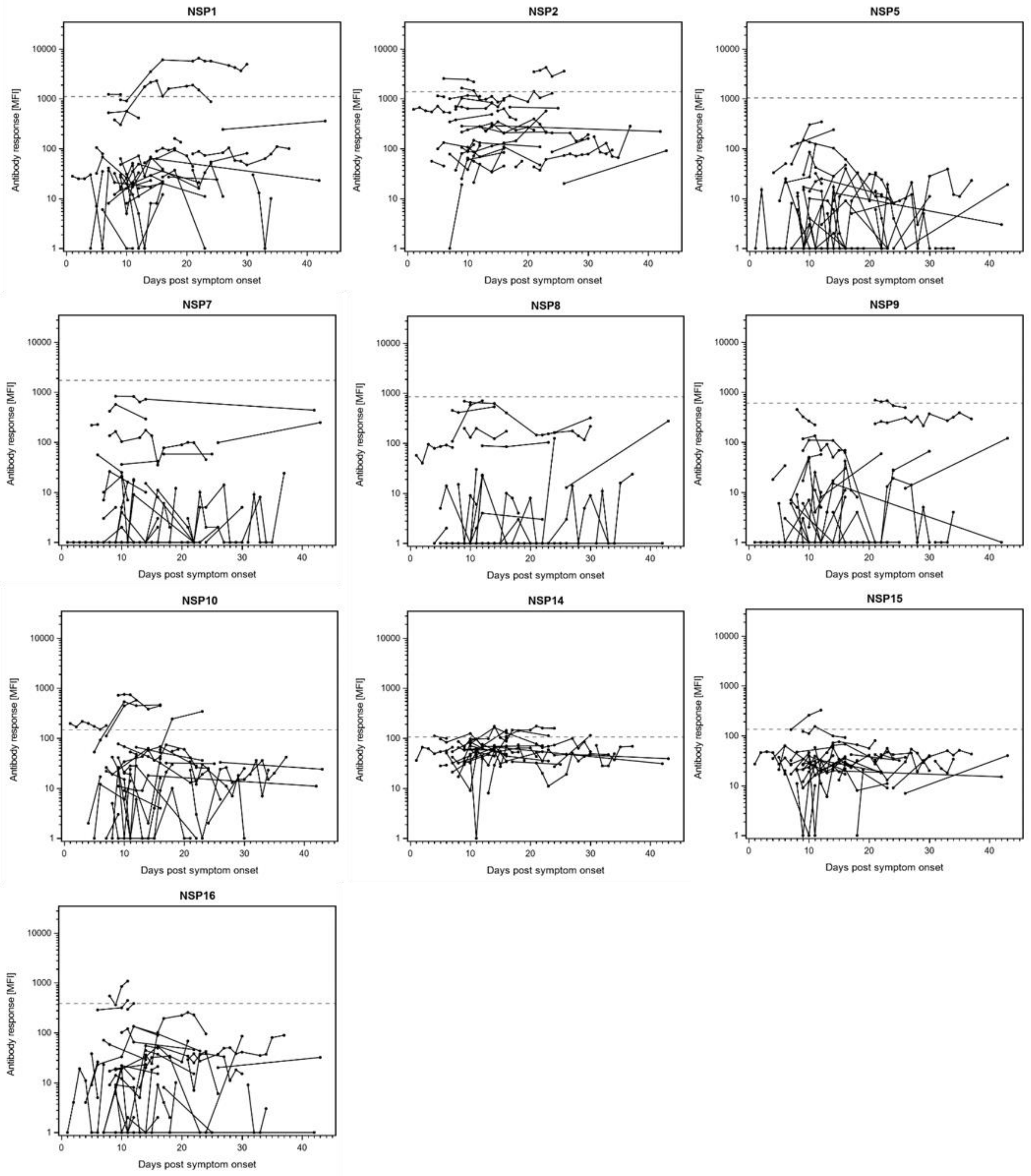


**Supplementary figure S3a: Antibody responses (IgM/IgA/IgG) to SARS-CoV-2 non-structural proteins in longitudinal samples of n=32 hospitalized Covid-19 patients.** The antibody response (MFI) is plotted against days post symptom onset. The dashed line indicates the antigen-specific cut-off defined as mean plus three times standard deviation in n=85 pre-pandemic controls.

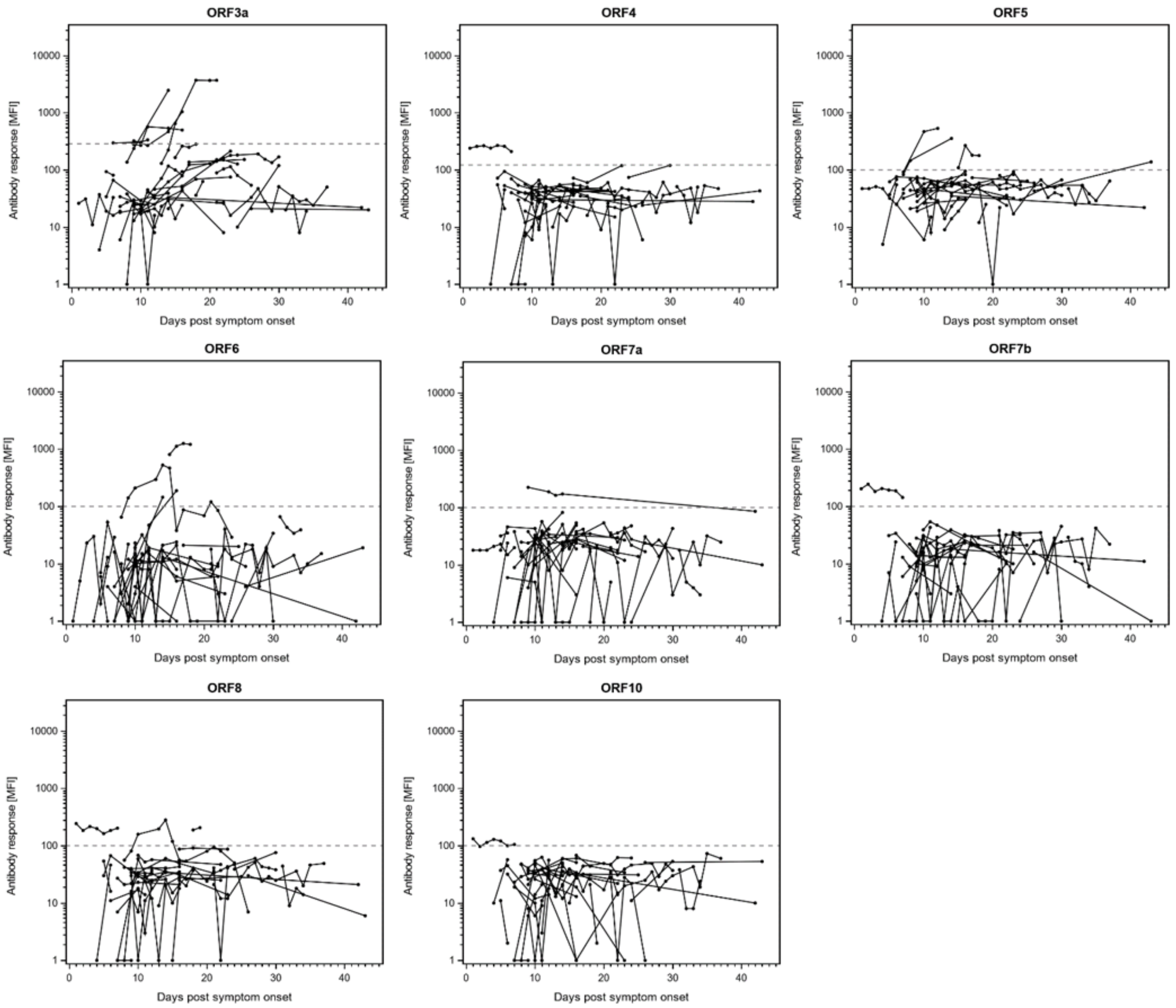


**Supplementary figure S3b: Antibody responses to SARS-CoV-2 ORF proteins in longitudinal samples of n=32 hospitalized Covid-19 patients.** The antibody response (MFI) is plotted against days post symptom onset. The dashed line indicates the antigen-specific cut-off defined as mean plus three times standard deviation in n=85 pre-pandemic controls.

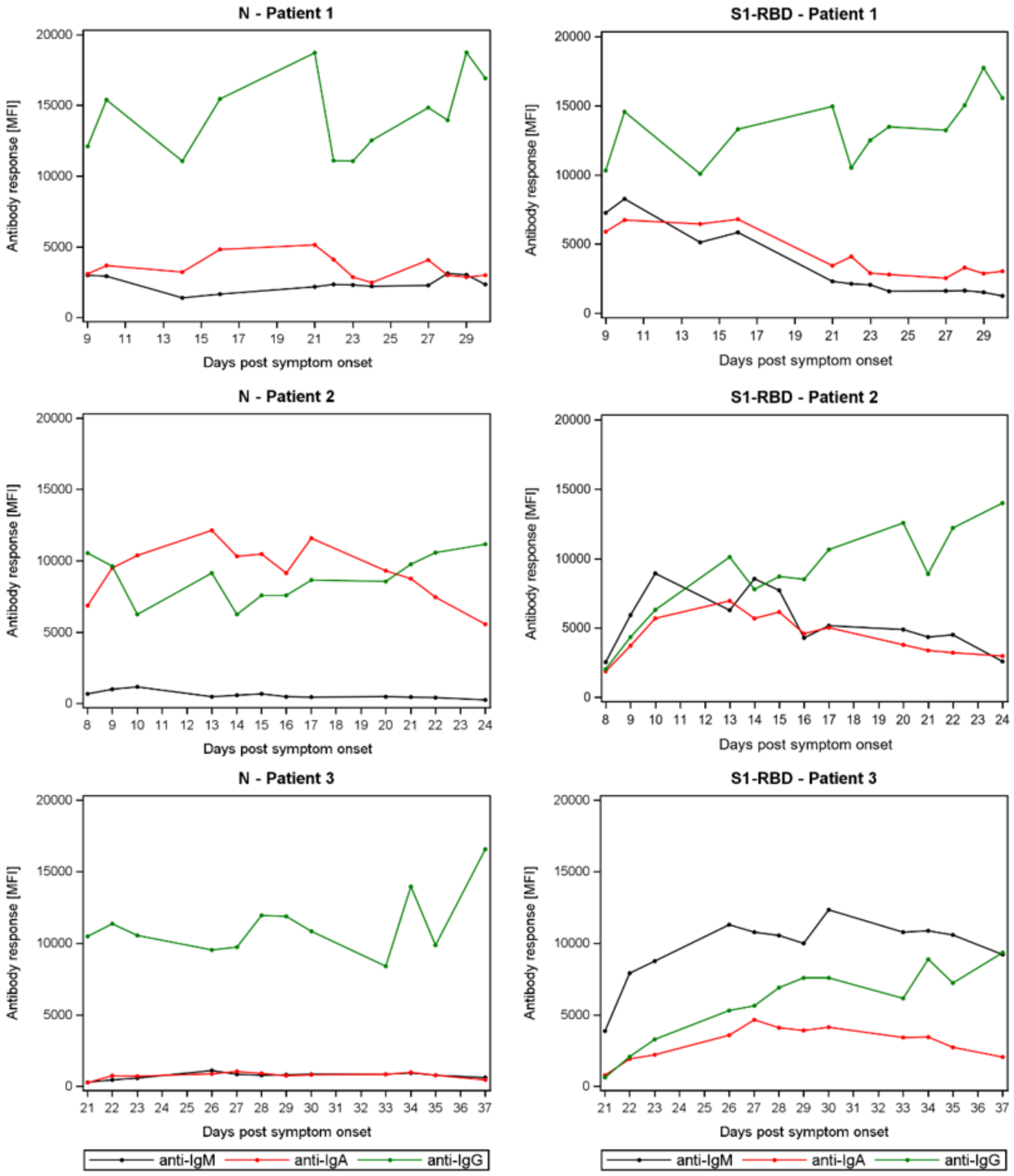


**Supplementary figure S4: Antibody response of different immunoglobulin isotypes (M, A, and G) to SARS-CoV-2 proteins N and S1-RBD in three Covid-19 patients with longitudinal samples.** The IgM, IgA, and IgG antibody response (MFI) is plotted against days post symptom onset.
